# To isolate, or not to isolate: a theoretical framework for disease control via contact tracing

**DOI:** 10.1101/2020.05.26.20113340

**Authors:** Davin Lunz, Gregory Batt, Jakob Ruess

## Abstract

Contact tracing is an essential tool in the public health battle for epidemiological control of infectious diseases. Contact tracing via case-by-case interviews is effective when contacts are known and outbreaks are small. Smartphone applications that keep track of contacts between users offer the possibility to scale contact tracing to larger outbreaks with minimal notification delays. While the benefits of reduced delays are widely recognised, it is less well understood how to best implement the tracing and notification protocol. The application will detect a multitude of contacts encountering an individual who later tests positive. Which of these contacts should be advised to self-isolate? The resolution hinges on an inherent trade-off: the more contacts notified, the greater the disease control, at the cost of more healthy individuals being instructed to self-isolate. In this study, based on a compartmental model tailored to the COVID-19 pandemic, we develop a framework to incorporate testing with limited resources coupled with a mechanistic description of digital contact tracing. Specifically, we employ a family of distributions characterising contact exposure and infection risk, and introduce a notification threshold that controls which level of exposure triggers notification. We detail how contact tracing can prevent disease outbreak, as a function of adoption rate, testing limitations, and other intervention methods such as social distancing and lockdown measures. We find an optimal notification threshold that balances the trade-off by minimising the number of healthy individuals instructed to self-isolate while preventing disease outbreak.

## Significance statement

Efficient contact tracing is expected to be of major importance for maintaining control of the COVID-19 pandemic. However, successful deployment demands a minimal burden on the general public. Previous studies have modeled the role of contact tracing, but have not addressed how to balance these two competing needs. We propose a modeling framework that captures contact heterogeneity. This allows contact prioritisation: contacts are only notified if they were acutely exposed to individuals who eventually tested positive. The framework thus allows us to address the delicate balance of preventing disease spread while minimising the social and economic burdens of quarantine, while being directly adaptable to different contact tracing implementations.

## Introduction

The COVID-19 pandemic has seen worldwide outbreaks, resulting in over five million validated cases of infection, and hundreds of thousands of deaths (at the time of writing). The enormous strain on healthcare infrastructure has led numerous countries to deploy their entire arsenal of epidemiological control measures to limit the spread of the disease. Epidemiological modeling has become one of the most important tools to inform political decisions on which control measures to deploy in a given situation [3]. A variety of modeling approaches are useful for this purpose, including branching processes [15, 19], network models [16, 6], age-structured models [8], stochastic differential equations [5], individual-based simulations [12], and classical compartmental models [11].

While contact tracing remains among the most important tools for epidemiological control, its use has been limited to small outbreaks due to the significant human resources required to trace contacts of infected individuals [8, 20, 10]. Moreover, the human-based approach may fail to identify contacts not personally known to an infected individual, which is especially relevant for highly infectious respiratory diseases as opposed to sexually transmitted diseases. Smartphone applications offer the possibility to overcome both the bottleneck and identification failure by making contact tracing scalable to larger outbreaks such as the present COVID-19 pandemic [7]. Compartmental models with contact tracing have been useful tools in modeling disease dynamics of HIV [14], Ebola [4], and even COVID-19 [22]. However, several aspects of these models severely limit their applicability.

On the one hand, the proportion of infected individuals removed due to contact traces is assumed to be independent of the outbreak size, which is only well justified when the outbreak remains relatively small. On the other hand, the removal of infected contacts is assumed to be proportional to both the number of traceable infected individuals and the number of infected contacts. Since this product is infinitesimally small compared to the disease dynamics terms at the disease-free equilibrium (DFE), additional underlying structure describing individual interactions needs to be assumed for the contact tracing terms to offer information on controlling the disease in the early (or late) stages of an outbreak [6, 19]. Thus, these models can only reliably describe outbreak attenuation for small epidemics, but cannot shed light on how contact tracing influences the initial outbreak, large disease outbreaks, or late-stage epidemics without significant added complexity.

One class of age-structured models [8] captures the impact of contact tracing at the disease-free equilibrium, however, several limitations persist. An exogenous contact tracing “efficacy” is prescribed a priori (the proportion of infections that are ultimately contact traced), which conceals the dependence of contact tracing on factors such as social intervention measures and contact tracing participation, while assuming that contact tracing efficacy is independent of the outbreak dynamics. Moreover, testing and contact tracing are assumed to be unrelated processes, which can lead to “substantial” errors [8].

Perhaps the most crucial ingredient absent in all of the aforementioned models is contact heterogeneity: contact is assumed either infectious or non-infectious. However, real-world contact is characterised by a spectrum of exposure levels, as would be detected by a digital contact tracing application [2, 1]. Neglecting this heterogeneity means that these models offer only limited guidance in prioritising which contacts to quarantine when dozens of contacts have encountered an infected individual.

In this paper, based on a compartmental model of COVID- 19, we develop a modeling framework that incorporates testing with limited capacities and a detailed mechanistic description of contact tracing. We capture contact heterogeneity by a generic contact exposure distribution, which describes the number of contacts encountered at different levels of *exposure*, typically accounting for the proximity and the duration of contact, and we associate to each exposure level a probability of infection. All contacts of positively diagnosed individuals that had an exposure greater than a controllable threshold are instructed to self-isolate. We establish how, from this setting, the total number of notified contacts and the proportion of those who contracted the disease from the traced individual, may be calculated from the epidemic state in combination with the contact exposure distribution. The tracing and isolation of these contacts is a dominant contribution to the disease dynamics near the DFE. In contrast to previous studies, the contact tracing process depends on both the testing and the epidemic state. Thus, our analysis provides precise quantification of the conditions required to prevent disease outbreak (or resurgence) when including contact tracing, as well as being able to inform policy when the disease is widespread within the population, without assuming additional population structure.

Importantly, both the contact exposure distribution as well as the corresponding infection probability may be freely specified in the framework. This allows the underlying epidemiological model to be customised with contact and infectiousness data of different locations or technology implementations.

We use our framework to investigate how saturated testing capacities and the resulting consequences for contact tracing lead to an acceleration of the epidemic during an early phase. We study how the basic reproduction number (the average number of secondary infections) determines the social intervention measures and adoption fraction required to prevent a disease outbreak. Combining these insights, we uncover an optimal notification threshold that minimises the numbers of individuals that need to self-isolate, while preventing disease outbreak. We deduce that a rise in social intervention measures or contact tracing adoption allows for a higher notification threshold, and hence fewer healthy individuals are unnecessarily advised to quarantine.

## Results

### Disease dynamics, testing and contact tracing framework

We model the disease dynamics via a compartmental model tailored to COVID-19 infection progression (Figure 1 left), while noting that our approach is directly adaptable to other infectious disease models. The model structure has been chosen to match previously reported key characteristics of COVID-19 such as a large fraction of presymptomatic infections [9, 17] and a non-negligible proportion of cases that remain completely asymptomatic and are unlikely to be identified [18]. Testing is performed on symptomatic individuals (compartment *I*): upon testing positive for the disease, a tested case is removed (compartment *R*, either recovered, isolated, or deceased, so assumed not to be infecting others). Thus, the base removal rate *γ_I_* is increased by the testing rate *τ*, which depends on the typical test rate *τ*_0_ and the maximal number of tests performed per unit time *τ_∞_* [24]. Note that these are *positive* testing rates, counting only the fraction of tests that result in positive diagnoses.

Central to the model is the contact tracing mechanism, which accounts for different exposure levels *e* when encountering contacts. The exposure level typically depends on the contact duration and proximity [1, 2]. For example, for diffusive virus transmission, the duration divided by the square of the distance provides an exposure measure. However, our approach is agnostic to the specific definition of exposure employed by any contact tracing application. We denote the distribution of contacts at an exposure level *e* by *ρ_c_*(*e*), and as sociate to each exposure level a probability of infection *p_i_(e)*. It is then natural to introduce a notification threshold, *u_n_*, such that only contacts exposed in excess of *u_n_* are advised to self-isolate.

**Figure 1:**
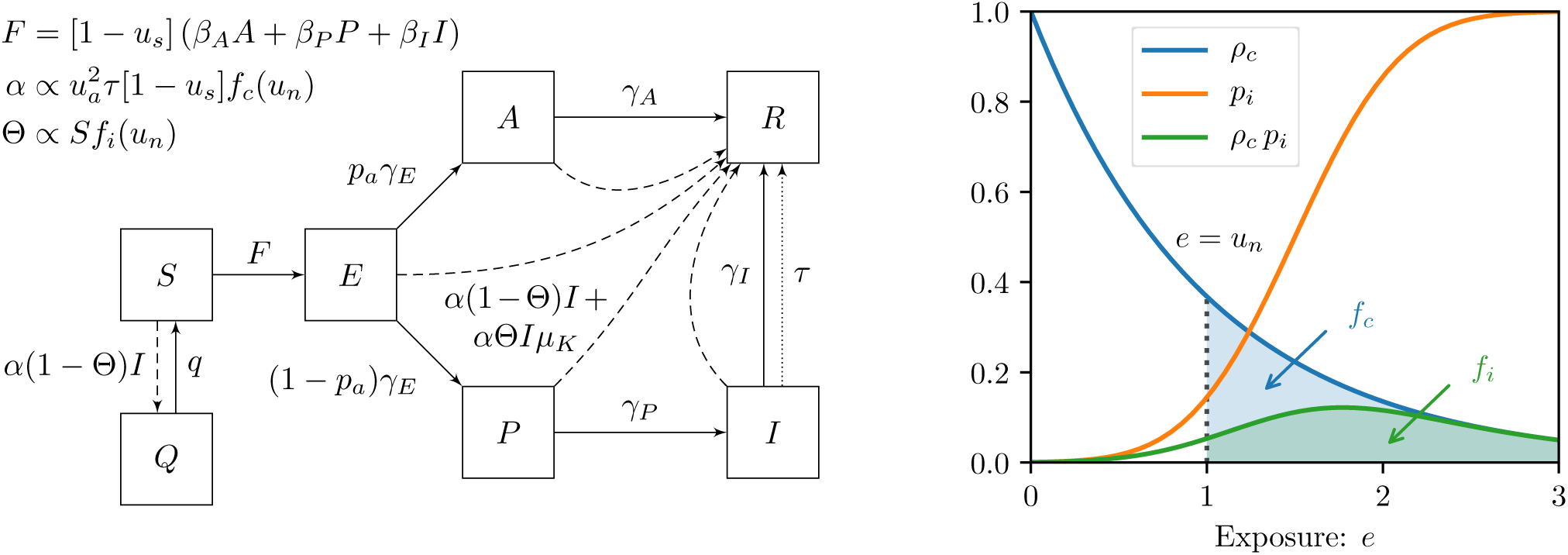
Disease dynamics and contact tracing framework. (left) Upon infection, susceptible individuals (*S*) enter a latent exposed stage (*E*) when the disease incubates before infectiousness. Upon becoming infectious, a proportion *p_a_* of the population remain asymptomatic (*A*), while the remainder pass through a presymptomatic stage (*P*) before becoming symptomatic (*I*). Infectious individuals are removed (*R*) through recovery/isolation (rates *γ_A_* and *γ_I_*), isolation after testing (dotted arrow, rate *τ*), or isolation as a consequence of contact tracing (dashed arrows, rate *α*(1 – Θ)*I* + *α*Θ*I_μK_* for *K* ∈ {*E; A; P; I*}). The total contact tracing rate *α* depends on the proportion of the population who adopt the contact tracing *u_a_*, the testing rate *τ*, the factor (1–*u_s_*(*t*)), representing the reduction of transmission rates due to social intervention measures and the notification threshold, *u_n_*, representing the minimal exposure required to notify traced contacts. Removal via contact tracing is partitioned into the fraction of contacts who were infected by the tested case Θ, and those who were not (1 – Θ), where Θ represents the tracing precision and depends on the susceptible proportion *S* and the notification threshold *u_n_*. Contact tracing also causes quarantine (*Q*) of susceptible individuals that had non-infectious contact with an infected case (dashed arrow at rate *α*(1 – Θ)*I*, return rate *q*). The force of infection, *F* depends on transmission rates *β_K_* and infection densities, and social intervention measures. (**right**) Heterogeneity in contacts is modeled with an exposure distribution, *ρ_c_*(*e*), and an infection probability function, *p_i_*(*e*), which describe the probability (density) that a given contact is of exposure *e* and the probability (mass) that a contact of given exposure *e* leads to an infection, respectively. The notification threshold (illustrated for *u_n_* = 1) affects the contact tracing rate and precision via the integral *f_c_* (blue and green shaded region) expressing the fraction of contacts notified, and the integral *f_i_* (green shaded region), which captures the proportion of contacts who had infectious contact with the tested case.

Equipping the disease dynamics with contact tracing removal (via the contact exposure *ρ_c_* and associated infection risk *p_i_*) allows us to derive expressions for the contact tracing rate, *α*, and the contact tracing precision (the proportion of notified contacts that were infected by the tested case), Θ. The notified contacts are separated into those infected by the tested case (true positives), and those not (false positives).

True positives are removed at the rate *α*Θ*I* from among the infectious and removed compartments in proportion to *μ_K_* for *K* ∈ {*E, A, P, I, R*}. The proportions *μ_K_* describe the probability of a traced contact having progressed (from their moment of infection) to compartment *K* by the time of contact tracing. True positives may reside in the *R* compartment, for example, those that, even though contracting the disease from the tested case, recovered sooner, or those that were false positives of a different contact trace. This is accounted for in our model by calculating the *μ_K_* distribution based on the stochastic progression of an individual through the disease stages depicted in Figure 1 left (see SI Appendix Section S1 B).

False positives are removed at the rate *α*(1 − Θ)*I* and are distributed uniformly among the entire population. Note that false positives may be infected if they contracted the disease from an individual other than the tested case. In this sense, true or false positiveness is with respect to a single tested individual.

Since the quantities *α*, Θ and *μ_K_* depend exclusively on the disease progression and contact tracing dynamics, they are derived directly from model analysis without introducing additional parameters or assumptions.

The full model may be written as an initial-value problem, comprising the following system of ODEs

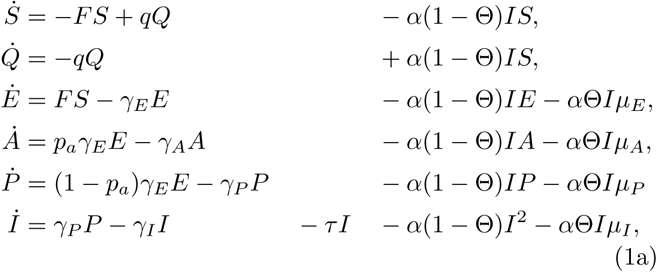

where

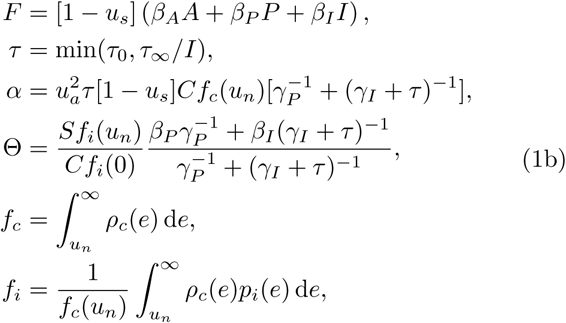

subject to the initial conditions

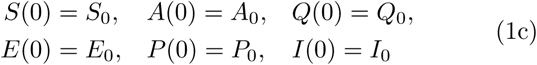

System (1) is derived from first principles in SI Appendix Section S1. The meaning of all parameters and controls is summarized in SI Appendix Tables 1 and 2. For the illustrations presented, we have adapted parameters from existing literature as outlined in SI Appendix Section S1 C. We highlight that the state variables describe population densities on the interval [0,1].

The framework incorporates three important control parameters: *u_a_* is the fraction of the population that adopt the contact tracing technology, *u_s_* describes the social intervention measures that reduce interpersonal contact, and *u_n_* is the notification threshold, describing the minimum exposure between a tested case and a contact that triggers a notification to self-isolate.

The tracing rate is proportional to the square of the adoption fraction 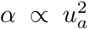, since only the fraction *u_a_* of tested cases, and only the fraction *u_a_* of traced contacts, participate in contact tracing. As opposed to previous work [7, 8], the contact tracing rate is proportional to, and is thus limited by, the testing rate and capacity and the social intervention measures *α* ∝ (1 − *u_s_*)*τ*. Similarly, contact tracing precision depends on the disease dynamics Θ ∝ *S*, with diminished precision in the presence of fewer susceptibles.

Armed with system (1), we establish how the crucial connection between testing, contact tracing, and isolation shapes the disease dynamics. Building upon these insights, we then demonstrate how analysis sheds light on the necessary intervention measures to prevent an epidemic. Finally, combining outbreak prevention with the interplay of testing, contact tracing, and isolation, we seek to prevent the epidemic while minimising unnecessary isolation notifications.

### The dual curse of limited testing resources

To demonstrate the intimate connection between testing, contact tracing, and isolation, we study their combined influence on an epidemic, that is, when disease outbreak is not prevented. In the first instance, we set the notification threshold *u_n_* = 0, where we notify all traced contacts, and assume that no social intervention measures are in place, *u_s_* = 0, focusing on the role of the maximal testing capacity *τ_∞_* for different adoption fractions *u_a_*. Since the contact tracing rate is proportional to the testing rate, *α* ∝ *τ*, as the disease spreads and infections rise in the population, testing capacity may become saturated *τ*_0_*I > τ_∞_*. When this happens, a smaller proportion of the symptomatic population are removed due to testing, the contact tracing removal suffers proportionally, and ultimately the epidemic accelerates.

To investigate the impact of limited testing capacities, we first compute the time *t* until 0.1% of the total population has been infected. For large enough testing capacity *τ_∞_*, the duration *t* of this early phase can be increased by nearly one month by case isolation alone *(u_a_* = 0), while for an adoption fraction of *u_a_* = 0.5, the epidemic can be slowed down by nearly two months (see Figure 2 left). Importantly, the dominant part of these gains are realized for *τ_∞_* on the order of the peak number of infections.

If the outbreak is not controlled in its early stages the disease invades the population. This is most drastic when testing capacity becomes saturated (see Figure 2 right). With increasing *τ_∞_*, the epidemic peak is delayed due to time gained during the early phase, and reduced in magnitude. Similarly, the total proportion of the population infected by the disease reduces significantly with increasing testing capacity *τ_∞_* and contact tracing participation *u_a_* (see SI Appendix Section S2C). This portfolio of improved outcomes is commonly referred to as “flattening the curve”.

We conclude that testing, contact tracing and isolation can buy valuable time to prepare the implementation of social intervention measures, while also reducing the peak and total strain on the healthcare infrastructure. However, the success of these measures critically depends on sufficient testing capacity *τ_∞_*. In particular, *τ_∞_* needs to be on the order of the infectious peak, which may be in excess of available testing resources. This highlights the need for broader social intervention measures that, in helping attenuate the epidemic, lighten the load on testing. When testing is overburdened the detrimental impact is two-fold: fewer individuals are diagnosed and self-isolate, which has the knock-on effect of curbing contact tracing.

**Figure 2:**
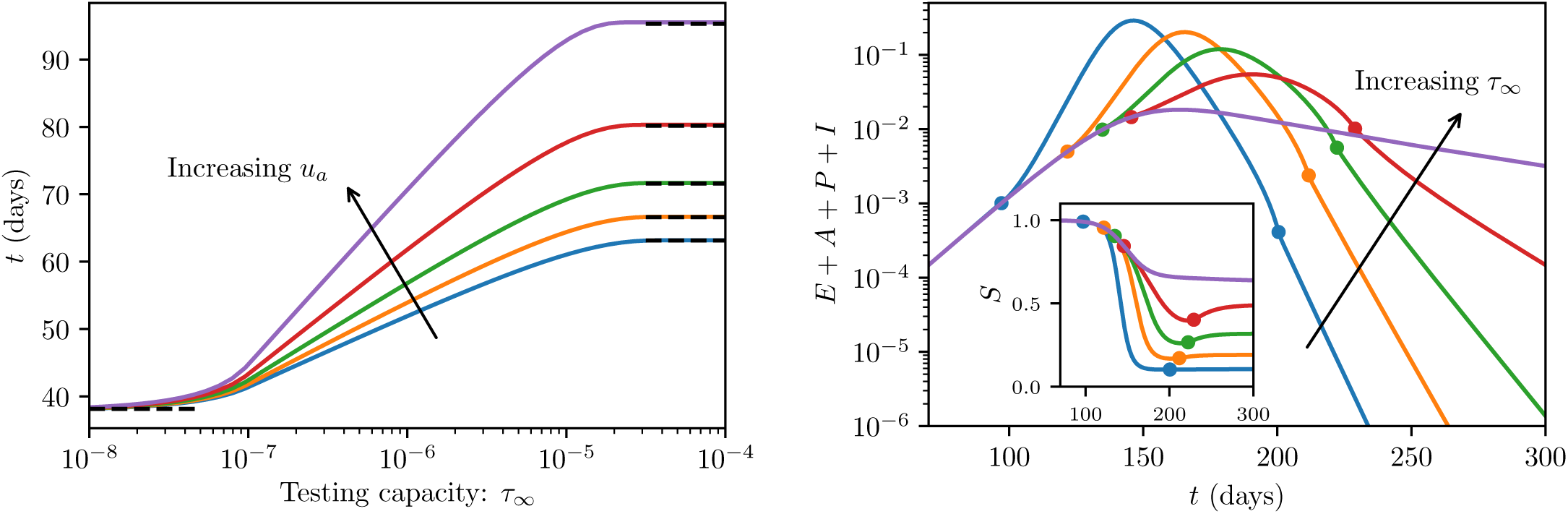
Flattening the curve. (left) Early-phase disease outcomes as a function of the maximum testing capacity *τ_∞_*. Colored curves correspond to an adoption fraction *u_a_ ∈* {0, 0.2,0.3, 0.4,0.5} and show the time *t* for 0.1% of the population to have been infected. Black dashed lines are asymptotic approximations (detailed in SI Appendix Section S2B). Increasing testing capacity and the adoption fraction decelerates the initial disease outbreak. **(right)** Long-term disease dynamics for *u_a_* = 0.4. Colored curves correspond to *τ_∞_* ∈ {1, 2, 5,10, 20} × 10^-4^ and show the total fraction of infected individuals *E* + *A* + *P* + *I*. If present, markers along the curves (of corresponding colour) show the time when the testing capacity saturates (smaller *t*) and desaturates (larger *t*). Increased testing capacity delays testing saturation, leading to smaller peak infection proportions. **[Inset]** Susceptible proportion *S* corresponding to the same simulation as the main plot. As testing desaturates the susceptible proportion rises. This is a result of a large number of healthy quarantined individuals *Q* returning to the susceptible pool.

The simulations demonstrate that the contact tracing resulted in many healthy individuals being unnecessarily quarantined (Figure 2 right inset). This motivates a more careful choice of notification threshold that balances sufficient disease control with the associated cost of excessive quarantining. First, we explore one approach to calculating sufficient disease control, and then we return to the question of prioritising contact isolation to lessen the quarantine cost.

### Contact tracing at the disease free equilibrium

Having demonstrated the descriptive power of the model when the disease is widespread, it is natural to ask whether the model can provide insight into the interventions necessary to prevent the epidemic. The basic reproduction number (the average number of secondary infections), denoted 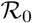, is a parameter that describes the disease outbreak threshold [23]: if 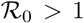 the disease-free equilibrium (DFE) is unstable, and the introduction of infected individuals leads to a disease outbreak, whereas if 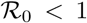 the introduction of a sufficiently small number of infected individuals does not lead to disease outbreak in the population. As opposed to earlier compartmental models [14, 4, 22], the contact tracing mechanism captures an 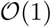 number of true-positive contacts, which are represented by terms that are linear with respect to *I* and thus their contribution is retained in the basic reproduction number 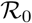 (see SI Appendix Section S2 A for details of the system regularisation via asymptotic analysis and SI Appendix Section S2 D for the calculation of the basic reproduction number). As a consequence, the influence of contact tracing on outbreak controllability can be analysed without simulating the full disease dynamics.

The basic reproduction number of the model (1) may be written in the form

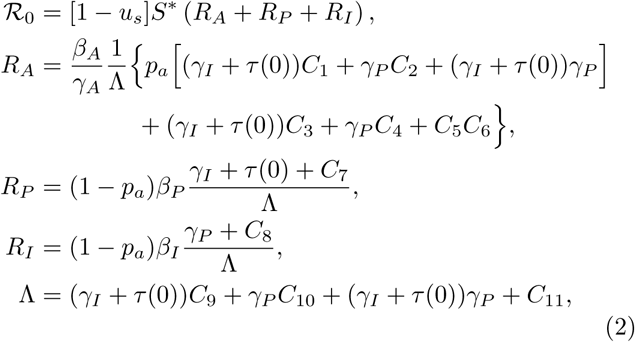

where *τ*(0) = sign(*τ*_∞_)*τ*_0_ (i.e. *τ*(0) = 0 if *τ_∞_* = 0) and the terms *C_i_* for *i* = 1,…, 11 represent (true positive) contact tracing contributions that depend on the problem parameters. Crucially, the quantities *C_i_* are proportional to the true positive removal rate *α*Θ and are thus linear with respect to social intervention measures (1 − *u_s_)* and the square of the adoption fraction 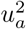. Therefore, in the absence of contact tracing, *u_a_ →* 0, these terms vanish, and the basic reproduction number reduces to the form

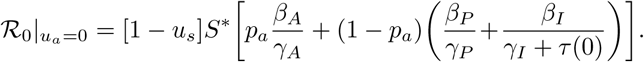

Since 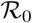 depends on the intervention parameters *u_s_*, *u_a_* and *u_n_*, we may explore outbreak prevention within a rich parameter space. Naturally, increasing social intervention measures *u_s_* and increasing the adoption fraction *u_a_* lead to a reduced number of secondary infections 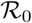 (see Figure 3 left). When 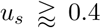 the value of 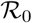 is below one and an outbreak is prevented based only on social distancing and case isolation. Contact tracing can reduce 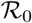 further but is not strictly necessary for outbreak prevention. Alternatively, if more than 80% of the population participate in contact tracing, an outbreak is prevented with no social intervention measures. For small adoption fractions *u_a_*, however, contact tracing is noticeably less effective. This observation reflects the dependence on the square of the adoption fraction 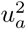, and highlights the importance of high levels of participation.

The basic reproduction number (2) exhibits a non-linear dependence on the proportion of susceptibles *S*\*. This is due to the factor of *S*\* in (2), as well as the contact tracing precision Θ depending on *S\**, which is inherited by the contact tracing terms *C_i_* (see SI Appendix Section S2D). Neglecting the dependence of the tracing precision on the susceptible population leads to an underestimation of the intervention measures required to prevent disease outbreak, which becomes significant when there is notable immunity in the population (see Figure 3 right). This is particularly relevant when lockdown measures are lifted after the first wave of the epidemic has passed.

**Figure 3:**
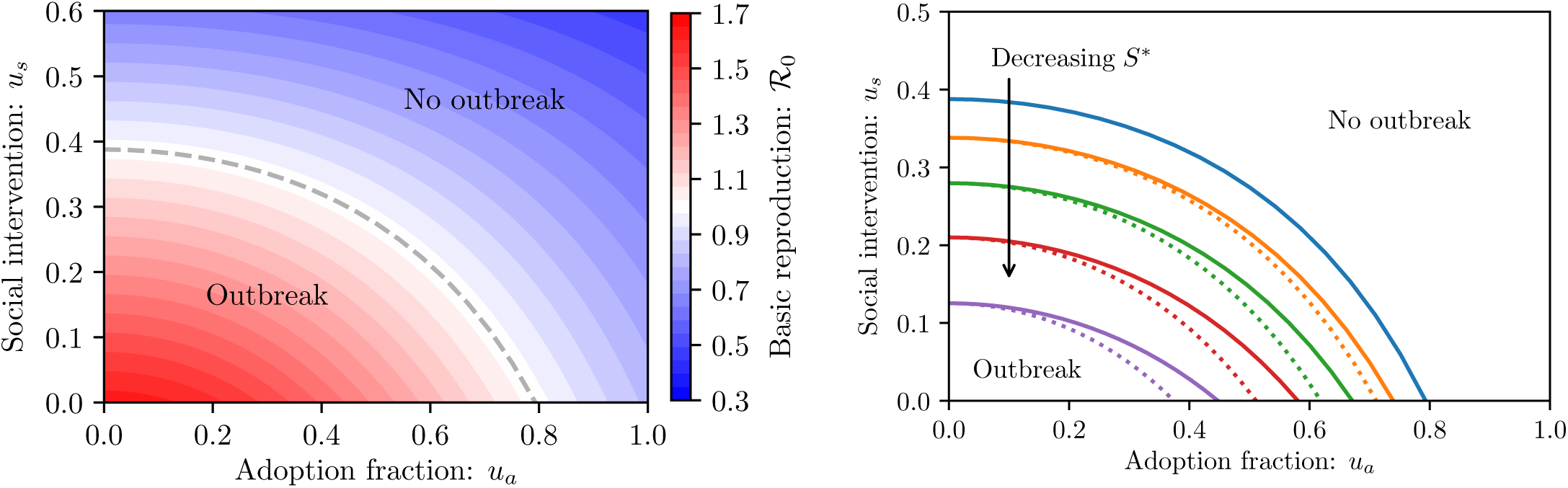
Conditions for outbreak prevention via 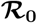 analysis. (left) Basic reproduction number 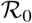 for *S\** = 1 and *u_n_* = 0 as a function of adoption fraction *u_a_* and social interventions *u_s_*. The dashed curve shows the 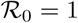 level set, which is the intervention threshold separating an outbreak from no outbreak. Increasing social intervention *u_s_* or contact tracing adoption *u_a_* increases disease control. **(right)** Intervention thresholds for *S*\* ∈ {1,0.925,0.85,0.775,0.7} and *u_n_* = 0. Dotted curves show the level sets 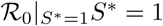 and correspond to neglecting the dependence of the contact tracing efficiency on *S*\*. Previous contact tracing descriptions do not account for the susceptible proportion of the population, and thus underestimate the necessary disease control.

We conclude that, to accurately analyze outbreak prevention, it is essential to capture the influence of contact tracing at the DFE, while accounting for the intricate interplay between the contact tracing, the population immunity, and social intervention measures. Having studied the disease prevention, we proceed to explore how to minimise unnecessary quarantining, while still preventing disease outbreak.

### Optimal digital contact tracing

To complete the characterisation of the disease outbreak and dynamics in the *(u_a_*, *u_s_*, *u_n_*)-space, we explore the impact of the final control parameter: the notification threshold *u_n_*. It might be tempting to minimise the basic reproduction number 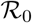, which would be achieved by simply setting *u_n_* = 0 to notify all contacts and avoid missing any true positive traced contacts. However, for small *u_n_*, there is no noticeable change in the 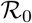 value (Figure 4 left, inset), as low-exposure encounters are unlikely to lead to an infection. Furthermore, there is a social and economic cost in requiring people to self-isolate unnecessarily, which we quantify by integrating the susceptible proportion of the population in quarantine up to some time 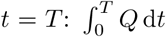. A higher notification threshold *u_n_* can achieve similar reductions in 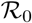 while lowering this quarantine cost (Figure 4 left). This suggests that we can find an optimal notification threshold that minimises the quarantine cost while still preventing an outbreak. The minimum reflects the trade-off inherent in the choice of notification threshold: *u_n_* is to be set low enough so that contact tracing occurs at a sufficiently high rate *α* to control the disease, but high enough so that tracing precision Θ ensures not too many susceptibles are quarantined (see SI Appendix Section S3 B).

**Figure 4:**
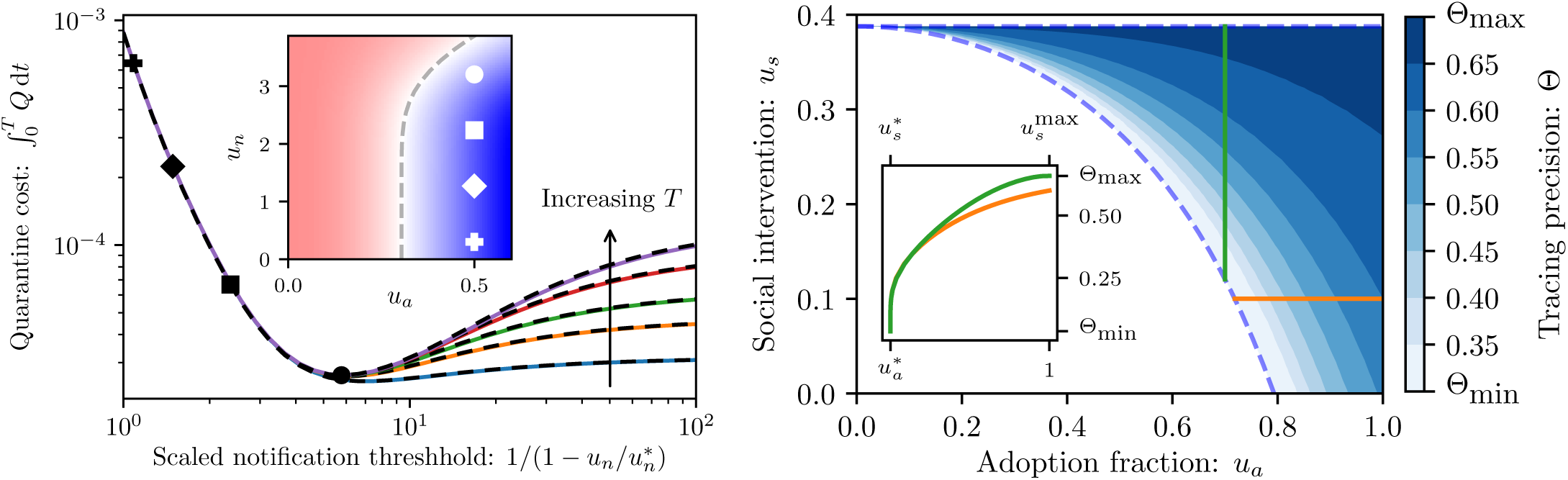
To isolate or not to isolate. (left)? Quarantine cost 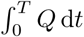 as a function of notification threshold u *u_a_* = 0.5, *u_s_* = 0.35, and *T* ∈ {1,1.5, 2, 3, 4} years. The notification threshold axis is mapped from the interval [0, 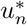] to [1, ∞] so as to spread out the region near criticality 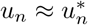 to illustrate the minimum cost (with 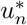 denoting the notification threshold for which 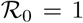). Each marked *u_n_* value corresponds to an identical mark in the inset. As *u_n_* increases from *u_n_* = 0 to the value at which the minimum is obtained, the quarantine cost improves by nearly two orders of magnitude while there is little variation in 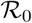. Black dashed curves show the asymptotic approximation derived in SI Appendix Section S2 B. **[Inset]** Basic reproduction number 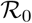 for *u_s_* = 0.35 as a function of *u_a_* and *u_n_*. The dashed line shows the 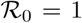 level set. **(right)** Optimal contact tracing precision Θ (i.e. the precision associated with the optimal notification threshold), as a function of social intervention measures *u_s_* and the adoption faction *u_a_*, within the region where an outbreak is controllable for sufficiently small *u_n_* but not controllable for arbitrarily large *u_n_*. At the lower 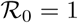 boundary only *u_n_* =0 prevents the epidemic, which corresponds to the minimal precision Θ_min_. Beyond the boundary there is a rapid increase in the optimal *u_n_* and thus a rapid increase in precision. In the vicinity of the upper boundary the optimal *u_n_* diverges, corresponding to the precision converging to Θ_max_. **[Inset]** One-dimensional slices of the tracing precision for fixed *u_a_* (green) and fixed *u_s_* (orange). We denote by 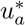 and 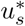 the critical parameter values for which 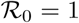 when all else is fixed, while 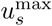 the upper boundary of the region, beyond which there is no outbreak even in the absence of contact tracing.

In going beyond the static calculations at the DFE, we must choose an appropriate time horizon *T*. Since the cost function admits a minimum that is fairly insensitive to the time horizon (Figure 4 left), we choose *T* = ∞, and seek the optimal notification threshold: *u_n_* that minimises the quarantine cost while preventing an outbreak. Since the units of *u_n_* match exposure, which are implementation-dependent, we choose to focus on the associated contact tracing precision Θ, expressing the fraction of true positive traces. Aiming to prevent disease outbreak via contact tracing, we focus on the region of parameter space where contact tracing is necessary and able to prevent disease outbreak (the blue shaded region in Figure 4 right).

The optimal tracing precision Θ increases with both increasing *u_a_* as well as increasing *u_s_*, from which we deduce that increasing social intervention measures and contact tracing participation allows for less aggressive notification. Since we expect many more low-exposure contacts to be encountered than high-exposure contacts, even a small increase in *u_n_* can mean a significant increase in Θ, accompanied by a reduction in unnecessary quarantining. For example, the optimal notification threshold in Figure 4 left corresponds to notifying approximately 4% of detected contacts, at a precision of Θ ≈ 0.61 (meaning 61% of notified contacts were infected by the traced case) while still preventing an outbreak 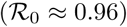. In comparison, for *u_n_* = 0 all contacts are notified, and thus 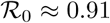, but contact tracing has a precision of only Θ ≈ 0.04. Unnecessarily quarantining 96% of notified contacts is an enormous social and economic burden that may undermine contact tracing acceptance. The associated reduction in the quarantine cost is nearly two orders of magnitude (compared with *u_n_* =0).

In summary, the model structure allows the notification threshold to be tuned, allowing a balance between aggressive contact tracing (low *u_n_* ensuring high disease control) and precise contact tracing (high *u_n_*, notifying contacts more likely to have been infected).

## Discussion

Strict social distancing policies, enforced to contain the COVID-19 outbreak, will have to be relaxed to prevent excessive damage to society and the economy. However, this must be achieved while avoiding a resurgent disease outbreak [21]. Contact tracing is one of the key measures for reducing the risk of subsequent epidemic waves, while allowing social distancing measures to be eased. Contact tracing needs to be fast and scalable to effectively disrupt infection chains (see [7] and SI Appendix Section S3D). Digital contact tracing via smartphone applications makes this possible, but raises the question of which among the many detected contacts to notify [13]. This problem hinges on an inherent trade-off: the lower the notification threshold, the greater the disease control, but the more healthy individuals sent unnecessarily to quarantine.

In this work, we embedded a compartmental model for COVID-19 disease progression, in a modeling framework that captures testing and digital contact tracing. Our results indicate that saturated testing capacities, and the consequence of less effective contact tracing, lead to an acceleration of the epidemic and more severe long-term outcomes. This suggests that the model can serve as a basis for quantitative studies on the role of limited PCR-testing during the spread of COVID- 19.

Our model introduces a mechanistic description of contact tracing in a compartmental model, which, guided by an asymptotic analysis, makes a dominant contribution at the disease free equilibrium and thus features in the basic reproduction number. This allowed us to derive contact tracing precision from the underlying process rather than prescribing an exogenous efficacy a priori [8, 7]. We investigated outbreak mitigation and prevention as a function of social intervention measures, contact tracing adoption (Figure 3 left), initial population immunity (Figure 3 right), testing rates (SI Appendix Section S3 A) and capacities (Figure 2), delays in contact notification (SI Appendix Section S3 D), and the notification threshold (Figure 4 left). The complex interplay in this high-dimensional parameter space highlights the practical challenge of achieving disease control. We emphasise that, when the outbreak is not preventable, contact tracing remains an important public safety measure: in many circumstances, every person who signs up to the contact tracing application saves another from infection (see SI Appendix Section S3 C).

The objective of our study was primarily to establish a comprehensive contact tracing modeling framework that can be adapted to a wide variety of models. Therefore, we did not present results for different parametrizations of the disease dynamics, which are expected to vary regionally and over time, but chose to focus on one set of values reasonable for COVID-19. While we have explored only constant control parameters *u_s_*, *u_a_*, and *u_n_*, we emphasise that the framework allows them to vary in time.

Our results do not qualitatively depend on the precise shapes of *ρ_c_*(*e*) and *p_i_*(*e*) (see SI Appendix Section S3 E). Nevertheless, to adapt the framework to a specific locale, it is important to determine these distributions from real-world data: the exposure distribution may be obtained directly from contact tracing platforms, and the infection probability can be deduced from the contact data in combination with further virological and epidemiological studies.

We have shown that our formulation of digital contact tracing, based on the contact exposure distribution *ρ_c_*(*e*) and corresponding relative infection probabilities *p_i_*(*e*), exposes a non-trivial notification threshold for optimal contact notification. Investigating this optimum reveals how, with more stringent social distancing measures or more adoption of the smartphone application, the contact tracing can be tuned to notify fewer contacts while still preventing an epidemic (Figure 4 right). Importantly, this leads to an overall reduction in unnecessary quarantining. Threshold adjustment allows policy makers to achieve a balance between disease management on the one hand, and social and economic cost on the other hand. We expect that our framework, within which this trade-off can be efficiently studied, will contribute to the implementation of digital contact tracing as a central tool in the sustainable fight against communicable diseases.

## Data Availability

No external datasets are used in this work.

## References

[1] Decentralized privacy-preserving proximity tracing. Technical report, DP-3T Project, 4 2020.

[2] Guidelines 04/2020 on the use of location data and contact tracing tools in the context of the covid-19 outbreak. Technical report, European Data Protection Board, 4 2020.

[3] F. Brauer. Mathematical epidemiology is not an oxymoron. BMC Public Health, 9(1):S2, 2009.

[4] C. Browne, H. Gulbudak, and G. Webb. Modeling contact tracing in outbreaks with application to ebola. Journal of Theoretical Biology, 384:33–49, 2015.

[5] S. Clémençon, V. Chi Tran, and H. de Arazoza. A stochastic SIR model with contact-tracing: large population limits and statistical inference. Journal of Biological Dynamics, 2(4):392–414, 2008. PMID: 22876905.

[6] K. T. D. Eames and M. J. Keeling. Contact tracing and disease control. Proceedings of the Royal Society of London. Series B: Biological Sciences, 270(1533):2565-2571, 2003.

[7] L. Ferretti, C. Wymant, M. Kendall, L. Zhao, A. Nurtay, L. Abeler-Dörner, M. Parker, D. Bonsall, and C. Fraser. Quantifying sars-cov-2 transmission suggests epidemic control with digital contact tracing. Science, 2020.

[8] C. Fraser, S. Riley, R. M. Anderson, and N. M. Ferguson. Factors that make an infectious disease outbreak controllable. Proceedings of the National Academy of Sciences, 101(16):6146–6151, 2004.

[9] T. Ganyani, C. Kremer, D. Chen, A. Torneri, C. Faes, J. Wallinga, and N. Hens. Estimating the generation interval for covid-19 based on symptom onset data. medRxiv, 2020.

[10] J. Hellewell, S. Abbott, A. Gimma, N. I. Bosse, C. I. Jarvis, T. W. Russell, J. D. Munday, A. J. Kucharski, W. J. Edmunds, S. Funk, and R. M. Eggo. Feasibility of controlling covid-19 outbreaks by isolation of cases and contacts. The Lancet, 8(4):e488-e496, 2020.

[11] H. W. Hethcote. The mathematics of infectious diseases. SIAM Review, 42(4):599–653, 2000.

[12] R. Hinch, W. Probert, A. Nurtay, M. Kendall, C. Wymant, M. Hall, K. Lythgoe, A. Bulas Cruz, L. Zhao, A. Stewart, L. Ferretti, M. Parker, A. Meroueh, B. Mathias, S. Stevenson, D. Montero, J. Warren, N. K. Mather, A. F. Finkelstein, L. Abeler-Dörner, D. Bonsall, and C. Fraser. Effective configurations of a digital contact tracing app: A report to NHSX. Technical report, 4 2020.

[13] R. Hinch, W. Probert, A. Nurtay, M. Kendall, C. Wymant, M. Hall, K. Lythgoe, A. B. Cruz, L. Zhao, A. Stewart, L. Ferretti, M. Parker, A. Meroueh, B. Mathias, S. Stevenson, D. Montero, J. Warren, N. K. Mather, A. Finkelstein, L. Abeler-Dörner, D. Bonsall, and C. Fraser. Effective configurations of a digital contact tracing app: A report to nhsx. Technical report, 2020.

[14] J. M. Hyman, J. Li, and E. A. Stanley. Modeling the impact of random screening and contact tracing in reducing the spread of HIV. Mathematical Biosciences, 181(1):17–54, 2003.

[15] C. Jacob. Branching processes: their role in epidemiology. International Journal of Environmental Research and Public Health, 7(3):1186–1204, 2010.

[16] I. Z. Kiss, D. M. Green, and R. R. Kao. Disease contact tracing in random and clustered networks. Proceedings of the Royal Society B: Biological Sciences, 272(1570):1407-1414, 2005.

[17] S. Ma, J. Zhang, M. Zeng, Q. Yun, W. Guo, Y. Zheng, S. Zhao, M. H. Wang, and Z. Yang. Epidemiological parameters of coronavirus disease 2019: A pooled analysis of publicly reported individual data of 1155 cases from seven countries. medRxiv, 2020.

[18] K. Mizumoto, K. Kagaya, A. Zarebski, and G. Chowell. Estimating the asymptomatic proportion of 2019 novel coronavirus onboard the princess cruises ship. medRxiv, 2020.

[19] J. Müller, M. Kretzschmar, and K. Dietz. Contact tracing in stochastic and deterministic epidemic models. Mathematical Biosciences, 164(1):39–64, 2000.

[20] C. M. Peak, L. M. Childs, Y. H. Grad, and C. O. Buckee. Comparing nonpharmaceutical interventions for containing emerging epidemics. Proceedings of the National Academy of Sciences, 114(15):4023–4028, 2017.

[21] H. Salje, C. Tran Kiem, N. Lefrancq, N. Courtejoie, P. Bosetti, J. Paireau, A. Andronico, N. Hoze, J. Richet, C.-L. Dubost, Y. Le Strat, J. Lessler, D. Levy Bruhl, A. Fontanet, L. Opatowski, P.-Y. Boelle, and S. Cauchemez. Estimating the burden of sars-cov-2 in france. medRxiv, 2020.

[22] B. Tang, X. Wang, Q. Li, N. L. Bragazzi, S. Tang, Y. Xiao, and J. Wu. Estimation of the transmission risk of the 2019-ncov and its implication for public health interventions. Journal of Clinical Medicine, 9(2), 2020.

[23] P. van den Driessche and J. Watmough. Reproduction numbers and sub-threshold endemic equilibria for compartmental models of disease transmission. Mathematical Biosciences, 180(1):29–48, 2002.

[24] W. Wang. Backward bifurcation of an epidemic model with treatment. Mathematical biosciences, 201(1–2):58-71, 2006.

